# Comparative Efficacy and Safety of Poly (ADP-Ribose) Polymerase Inhibitors in Cancer: Systematic Review and Network Meta-Analysis

**DOI:** 10.1101/2025.03.11.25323491

**Authors:** Geqiong Xiao, Junwei Yan, Zhidong Cai, Menglu Wu, Ruolin Zhang, Xiaopeng Yang, Yiying Yao, Zhongkui Xiong, Mengfei Ye, Jian Zhang, Zheng Liu

**Affiliations:** Department of Oncology, Affiliated Hospital of Shaoxing University, Shaoxing, Zhejiang, China; Department of Blood Transfusion, Affiliated Hospital of Shaoxing University, Shaoxing, Zhejiang, China; Department of Pharmacology, School of Medicine, Shaoxing University, Shaoxing, Zhejiang, China; Department of Radiation Oncology, Shaoxing Second Hospital, Shaoxing, China; Department of Psychiatry, Shaoxing seventh people’s hospital, Mental Health Center, School of Medicine, Shaoxing University, Shaoxing, Zhejiang, China

**Keywords:** PARP inhibitors, Cancer, Efficacy, Safety, Network meta-analysis

## Abstract

**Background:** The clinical background of cancer patients is complex, and the efficacy and safety of different poly (ADP-ribose) polymerase inhibitors (PARPis) vary greatly. Therefore, we conducted a systematic review and network meta-analysis to compare the efficacy and safety of the six PARPis and systematically evaluated the differences in PARPis in the relevant clinical contexts.

**Methods:** Databases (PubMed, Cochrane, Embase, etc.) were searched from Jan 1, 1980 to Oct 20, 2023, for randomized clinical trials. The overall survival (OS), progression-free survival (PFS), and grade 3-5 adverse events (AEs) were the main outcomes and measures.

**Results:** Forty-seven trials targeting 17 300 patients comparing six different PARPis. Talazoparib was associated with the highest PFS (HR = 0.59, 95% CI: 0.52-0.68), accompanied by grade 3-5 AEs (HR = 4.96, 95% CI: 1.49-18.08). There were additional potential beneficial relationships in populations: homologous recombination deficiency (HRD) with PFS (HR = 0.34, 95% CI: 0.24-0.43); BRCA gene mutations with OS (HR = 0.79, 95% CI: 0.68-0.93) and PFS (HR = 0.30, 95% CI: 0.27-0.34); new diagnoses with OS (HR = 0.79, 95% CI: 0.61-0.97); and sensitivity to prior platinum therapy with PFS (HR = 0.43, 95% CI: 0.39-0.47).

**Conclusion:** PARPis are more effective in HRD, newly diagnosed or platinum-sensitive cancer populations, and talazoparib is recommended as the first choice.

## Introduction

Genomic instability is a hallmark of cancer,^1^ with approximately 1 in every 400–800 individuals harboring a BRCA1 or BRCA2 gene mutation that can result in homologous recombination deficiency (HRD).^2^ Excitingly, poly (ADP-ribose) polymerase inhibitors (PARPis) have been shown to induce synthetic lethality in tumor cells, primarily by inhibiting PAR acylation in HRD cancers.^3^

Recently, meta-analyses have shown that PARPis produces a better progression-free survival (PFS) in ovarian and breast cancer patients than does placebo.^4–10^ However, one study concluded that the PFS was not statistically significant in the non-HRD population.^11^ In addition, the benefit of PARPis in overall survival (OS) is also controversial.^5, 6, 9, 10^ Most studies found PARPis were more likely to cause grade 3–5 adverse events (AEs), but two studies did not find a difference.^9, 10^ These controversies may be due to the complex clinical background of cancer patients recruited by researchers.

Due to the limitations of traditional meta-analyses, these studies neither directly compared the efficacy and safety of the six PARPis nor systematically evaluated the differences in PARPis in the relevant clinical contexts. The inconsistent treatment outcomes of these drugs and the diversity of adverse events emphasize the need for more comprehensive guidelines for clinical decision-makers. Our network meta-analysis (NMA) effectively addressed the above issues. More specifically, we compared the effectiveness and safety of PARPis identified in randomized clinical trials on cancer, providing information for clinical decision-makers.

## Materials and Methods

### Eligibility Criteria

Our NMA was conducted in accordance with the Preferred Reporting Items for Systematic Reviews and Meta-Analyses extension statement for network meta-analyses^12^ (eTable 1). To ensure the transparency, reliability, and novelty of the research protocol, our study was registered with the Prospective Register of Systematic Reviews (CRD42023452136).

### Data Sources and Extraction

PubMed, Cochrane, Embase, ClinicalTrials.gov, and International Clinical Trials Registry Platform databases were searched for articles published from Jan 1, 1980, to Oct 20, 2023 (eTable 2). In addition, references to relevant articles or comments were manually checked to avoid omissions.

MLW and RLZ independently conducted literature searching and data extraction. All conflicts were resolved by discussion with a senior author (ZL). The extracted data items included characteristics of the studies, selection criteria, exclusion criteria, reported outcomes, hematologic serious AEs, and non-hematologic serious AEs (eTable 3). The effectiveness indicators were OS and PFS, and the safety indicators were grade 3–5 AEs.

### Risk of Bias Assessment

Two reviewers (XPY and YYY) used the Cochrane Handbook’s risk of bias assessment tool to assess the risk of bias of the eligible studies, and conflicts were resolved by discussion. Studies were evaluated from seven biases: random sequence generation, allocation concealment (selection bias), blinding of participants and personnel (performance bias), selective reporting (reporting bias), incomplete outcome data, blinding of outcome assessment (detection bias), and other biases.^13^ Based on the included studies, three categories were created: low risk, some concerns, and high risk. All conflicts were resolved by discussion with a senior author (ZL).

### Statistical Analysis

OS and PFS were the primary outcomes, and grade 3–5 AEs were the secondary outcomes. For OS and PFS, we used HRs with 95% CIs, and for grade 3–5 AEs, we used ORs with 95% CIs. The indirect comparison of each therapy was first conducted using OpenBUGS software (version 1.4.3) in a Bayesian framework. The evaluation of each result used a fixed effects consistency model, using three independent Markov chains to run 5000 burn-ins and 15 000 sample iterations with one step-size iteration per chain simultaneously. For AE analysis, our work was performed with R software (version 4.0.3) and the gemtc package (version 0.8–8) with 4 parallel Markov chains consisting of 50,000 samples after a 20 000-sample burn-in. Using a Bayesian approach, we also ranked each therapy according to its overall ranking probabilities.

We used random effects models to explain the heterogeneity between studies. We evaluate heterogeneity by using Cochran Q statistics and inconsistency statistics (I^2^). For I^2^, index values of less than 25% indicate low heterogeneity; 25% to 50% indicate medium heterogeneity; and greater than 50% indicate high heterogeneity.^14^

## Results

### Characteristics of the Included Studies

We extracted 3696 records from the databases, and after removing duplicate studies and excluding articles without full text, 142 studies remained. Through full text reviews, 47 studies met our eligibility criteria (eFigure 1). ^15–61^

**Figure 1.**
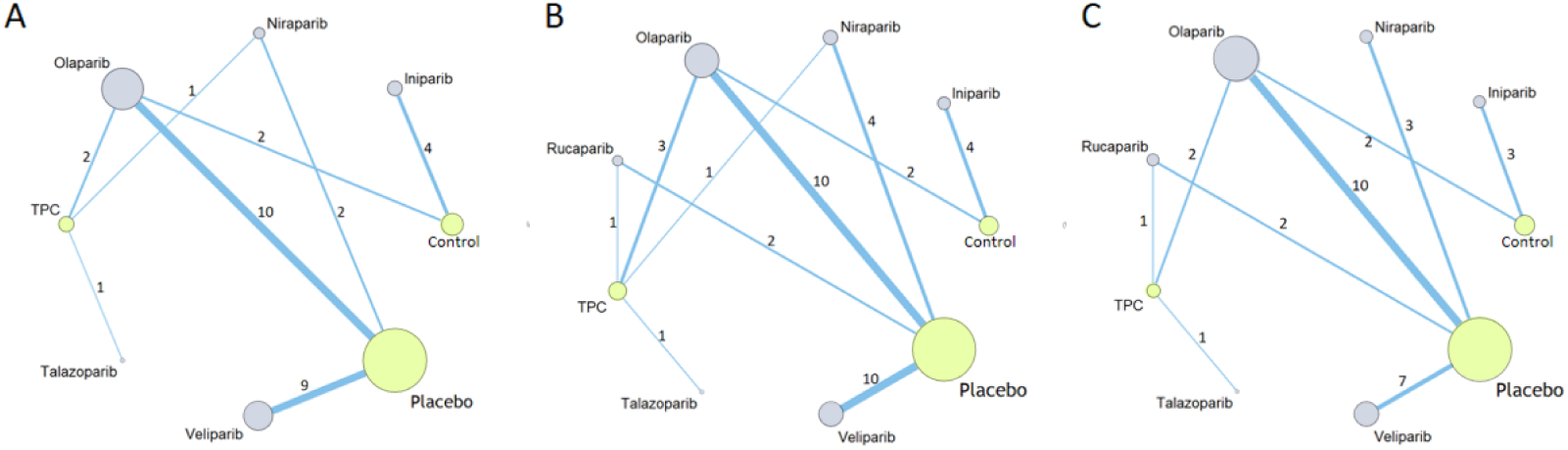
Networks for overall survival (A), progression-free survival (B), and grade 3–5 adverse events (C). Each circle corresponds to a treatment, and the area of the circle is proportional to the number of patients. Each line represents a direct comparison between two treatments, and the thickness of the line is proportional to the number of direct comparisons. Control represents that the patient no longer received other drugs (including placebos) except for the same treatment medication as the experimental group. TPC, treatment of physician’s choice.

Forty-seven trials targeting 17 300 patients compared nine treatments: iniparib, niraparib, rucaparib, talazoparib, veliparib, olaparib, treatment of physician’s choice (TPC), and placebo and control (no longer receiving other drugs except for the same treatment medication as the experimental group). Detailed information of all included studies can be found in eTable 3. The network plots of the research are depicted in Figure 1, and the assessment of risk of bias is shown in eFigure 2.

**Figure 2.**
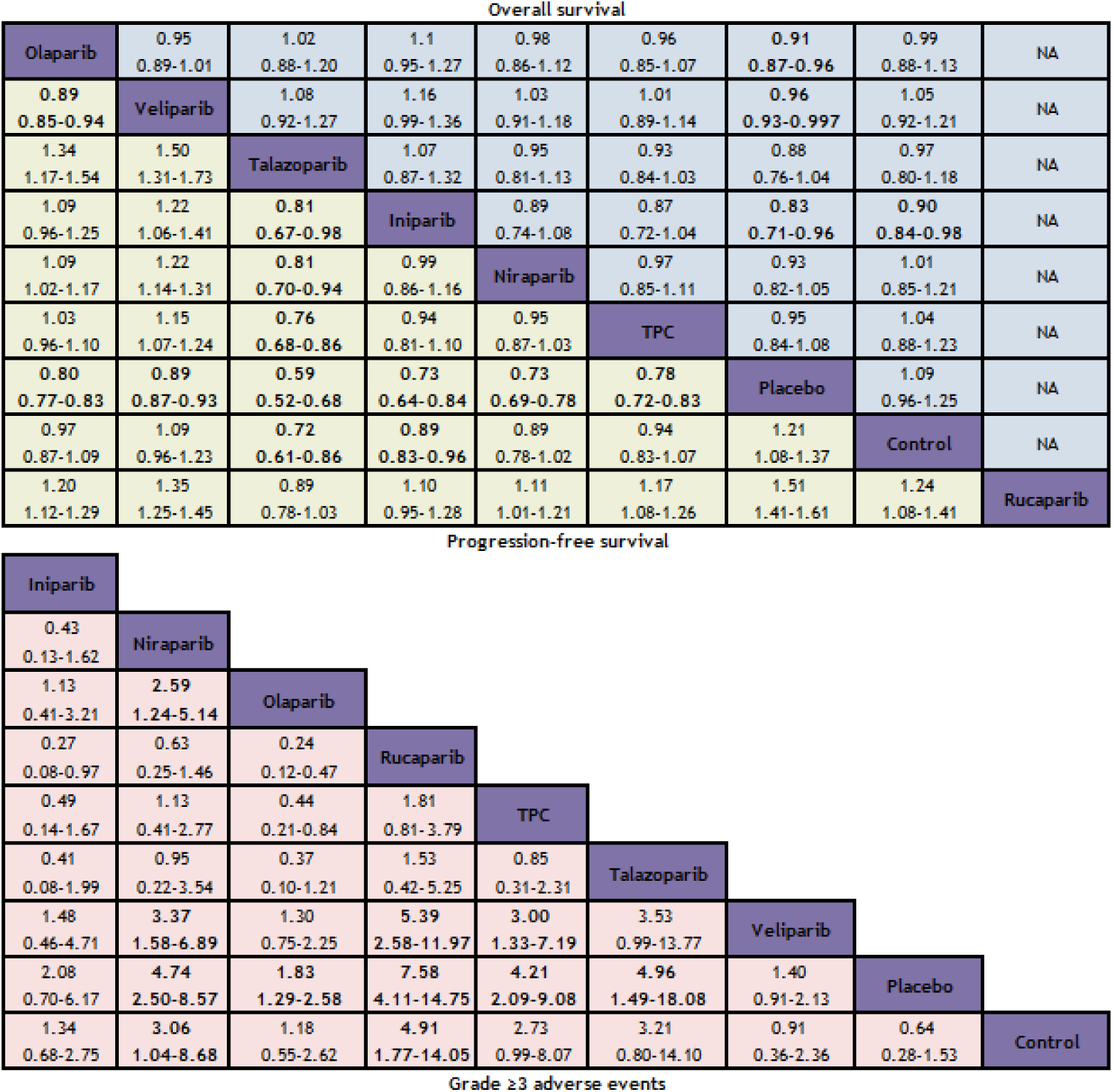
League table of unadjusted network meta-analyses for the risk of overall survival and progression-free survival (above), and grade 3–5 adverse events (below). The upper half of the grid shows odds ratios for OS; the lower half of the grid shows odds ratios for PFS. Each box represents the comparison of the row-defined treatment versus the column-defined treatment. Odds ratios of more than 1 favor the column-defining treatment and odds ratios of less than 1 favor the row-defining treatment. The comparison of the column-defining treatment versus the row-defining treatment is the reciprocal of the data shown. Control represents that the patient no longer received other drugs except for the same treatment medication as the experimental group. NA, not applicable. TPC, treatment of physician’s choice.

### Effectiveness

Regarding OS (Figure 2A), except for talazoparib (HR = 0.88, 95% CI: 0.76–1.04) and niraparib (HR = 0.93, 95% CI: 0.82–1.05), PARPis were more likely to produce a greater OS benefit than the placebo. Rucaparib lacked relevant clinical data on OS and was not included in the comparison. Of the PARPis, iniparib yielded the best OS benefit compared with placebo (HR = 0.83, 95% CI: 0.71–0.96). Following closely was olaparib (HR = 0.91, 95% CI: 0.87–0.96) and veliparib (HR = 0.96, 95% CI: 0.93–0.997).

PFS (Figure 2A) was broadly consistent with OS. PARPis were more likely to confer a greater PFS benefit than the placebo. Among the PARPis, talazoparib was associated with the longest PFS versus placebo (HR = 0.59, 95% CI: 0.52–0.68), followed by rucaparib (HR = 0.66, 95% CI: 0.62–0.71), iniparib (HR = 0.73, 95% CI: 0.64–0.84), niraparib (HR = 0.73, 95% CI: 0.69–0.78), and olaparib (HR = 0.80, 95% CI: 0.77–0.83). The least effective was veliparib (HR = 0.89, 95% CI: 0.87–0.93).

### Safety

The NMA included 9 treatments for grade 3–5 AEs (Figure 1C). Compared to placebo, rucaparib was most likely to cause grade 3–5 AEs (HR = 7.58, 95% CI: 4.11–14.75), followed by talazoparib (HR = 4.96, 95% CI: 1.49–18.08), niraparib (HR = 4.74, 95% CI: 2.50–8.57), and olaparib (HR = 1.83, 95% CI: 1.29–2.58). Furthermore, iniparib (HR = 2.08, 95% CI: 0.70–6.17) and veliparib (HR = 1.40, 95% CI: 0.91–2.13) showed no difference with the placebo (Figure 2B). Talazoparib exhibited the highest probability of causing anemia, veliparib was the most likely to cause neutropenia and leukopenia, and niraparib was the most likely to cause severe thrombocytopenia. The incidence of nonhematologic grade 3–5 AEs, such as fatigue, nausea, vomiting, diarrhea, and abdominal pain was a narrow incidence spectrum (Figure 3). Four studies.^43, 55–57^ on iniparib were not included in the analysis. Their incidence of adverse reactions differed significantly from the other studies, and were more likely to reflect adverse reactions caused by combination chemotherapy drugs.

**Figure 3.**
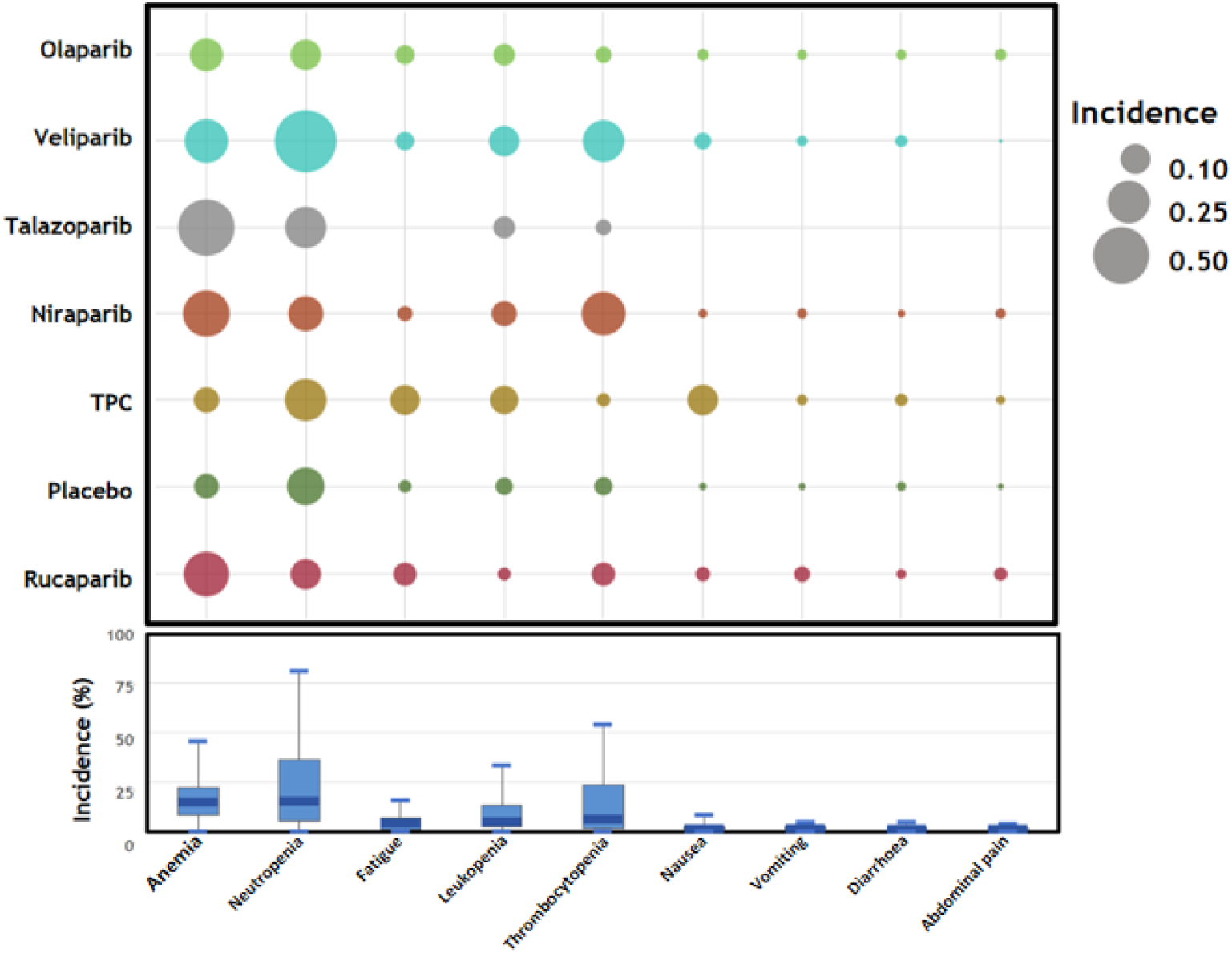
Incidence of grade 3–5 adverse events. TPC, treatment of physician’s choice.

### Ranking

The ranking analysis was based on Bayesian ranking profiles (Figure 4, eTable 4). Among the PARPis, talazoparib and rucaparib demonstrated significant therapeutic effects and high toxicity. Talazoparib ranked second in OS (49%) and first in PFS (93%), and ranked second in the number of grade 3–5 AEs (47%). Rucaparib ranked second in PFS (89%) and first for grade 3–5 AEs (65%). Iniparib showed clear efficacy and low toxicity, ranking first in OS (69%) and third in PFS (47%). Niraparib showed moderate efficacy and toxicity, and was fifth in OS (67%), fourth in PFS (90%), and fourth in grade 3–5 AEs (93%). Olaparib demonstrated moderate efficacy and low toxicity, and was ranked fourth in OS, sixth in PFS, and sixth in grade 3–5 AEs. Veliparib had low efficacy and toxicity, and was seventh in OS, eighth in PFS, and eighth in grade 3–5 AEs.

**Figure 4.**
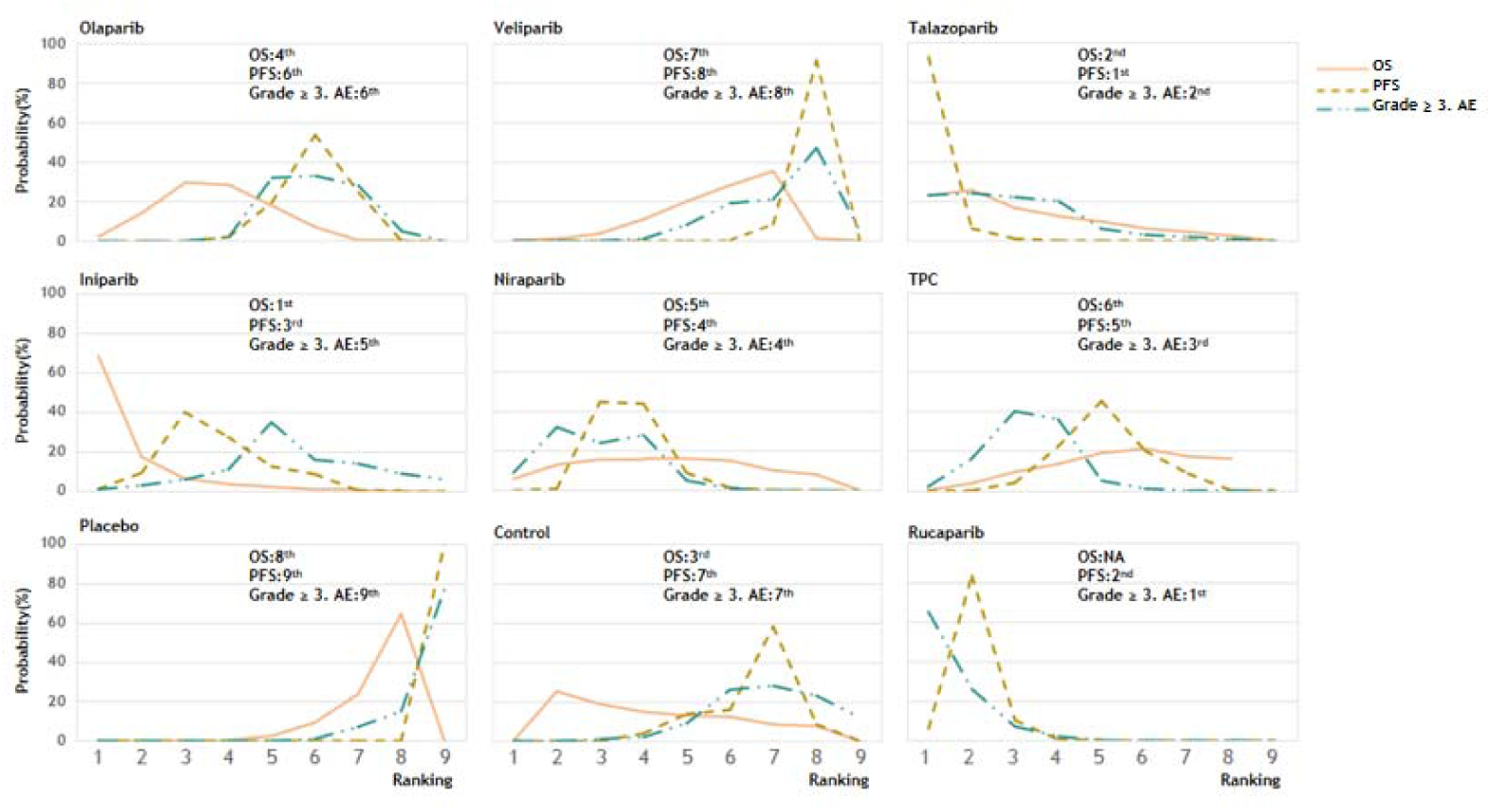
Bayesian ranking profiles of PARP inhibitors for efficacy and safety. Ranking plots indicate the probability of each inhibitor being ranked from first to last in OS, PFS, and grade 3–5 AEs. OS, overall survival; PFS, progression-free survival; AE, adverse event. Control represents that the patient no longer received other drugs except for the same treatment medication as the experimental group. TPC, treatment of physician’s choice. NA, not applicable.

### Subgroup Analysis

All subgroup analysis data is provided in Table 1.

**Table 1.**
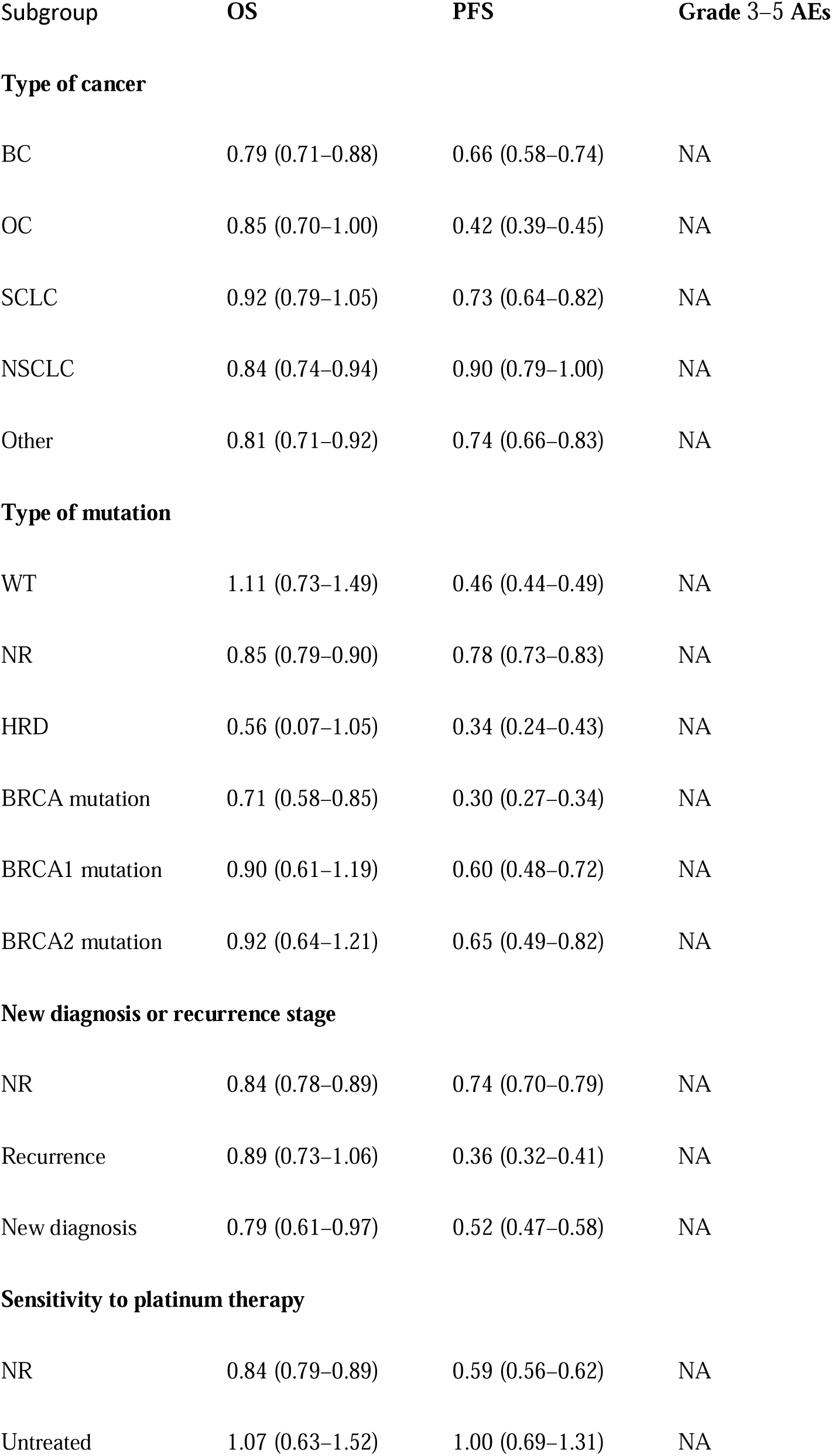
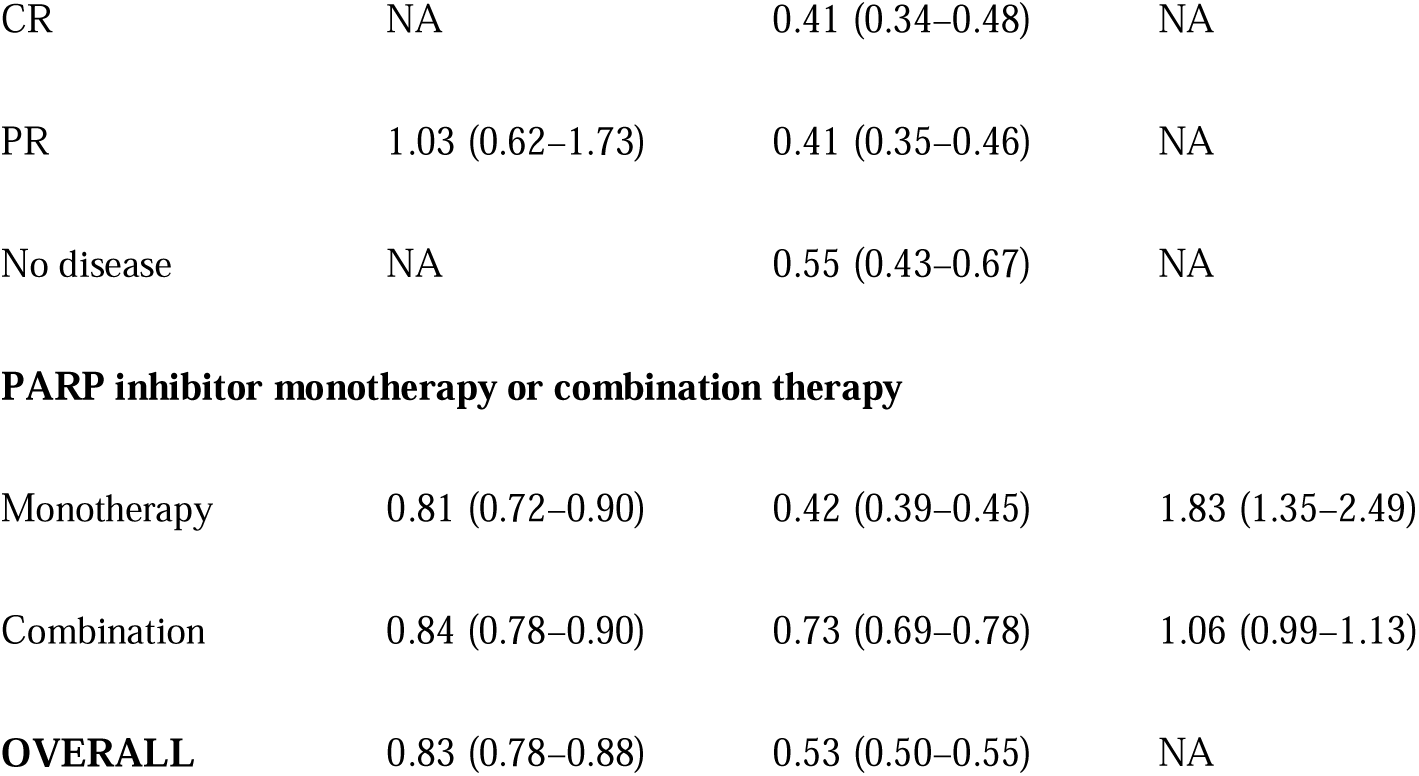
Subgroup of Meta-analysis Results.

### Types of Cancer

According to the types of cancer, patients were divided into the following five subgroups: breast cancer (BC), ovarian cancer (OC), non-small cell lung cancer (NSCLC), small cell lung cancer (SCLC), and other (urothelial carcinoma, pancreatic adenocarcinoma, melanoma, and pancreatic, colorectal, prostate, and gastric cancer). In the overall population, PARPis were associated with higher OS (HR = 0.83, 95% CI: 0.78–0.88) and PFS (HR = 0.53, 95% CI: 0.50–0.55) benefits compared to the control group.

Regarding OS, BC (HR = 0.79, 95% CI: 0.71–0.88), NSCLC (HR = 0.84, 95% CI: 0.74–0.94), and other (HR = 0.81, 95% CI: 0.71–0.92) had superior OSs compared to that of the control group. Regarding PFS, except for NSCLC (HR=0.90, 95% CI: 0.79–1.00), the other groups showed significant benefits compared to the control group.

### Types of Gene Mutations

Regarding OS, the BRCA gene mutation (HR = 0.79, 95% CI: 0.68–0.93), and not reported (NR) were superior to the control group. Regarding PFS, all subgroups demonstrated benefits. The BRCA gene mutation (HR = 0.30, 95% CI: 0.27–0.34) was associated with the highest benefit, followed by HRD (HR = 0.34, 95% CI: 0.24–0.43).

### New Diagnosis or Recurrence

Regarding OS, new diagnosis (HR = 0.79, 95% CI: 0.61–0.97) and NR were superior to the control group. Regarding PFS, all subgroups demonstrated good benefits, and those with recurrence (HR = 0.36, 95% CI: 0.32–0.41) demonstrated the highest benefits.

### Sensitivity to Platinum Therapy

Patients’ platinum therapy statuses were grouped as follows: not receiving platinum therapy (untreated), complete response (CR), partial response (PR), and no disease, NR.

Regarding OS, NR (HR = 0.84, 95% CI: 0.78–0.89) was superior to the control group. Regarding PFS, CR (HR = 0.41, 95% CI: 0.34–0.48), PR (HR = 0.41, 95% CI: 0.35–0.46), no disease (HR = 0.55, 95% CI: 0.43–0.67), and NR were superior to the control group.

### Monotherapy or Combination Therapy

Compared with combination therapy, PARPis monotherapy was similar to more effective as measured by OS (HR = 0.81, 95% CI: 0.72–0.90) and PFS (HR = 0.42, 95% CI: 0.39–0.45), but it was more likely to cause grade 3–5 AEs (HR = 1.83, 95% CI: 1.35–2.49).

### Risk of Bias

Among the 47 studies included, 2 were open label studies, and they showed that indicators with a higher risk than other indicators involved “selection bias (allocation concealment).” This indicated that the included studies had overall good internal validity (eFigure 2).

### Exploration for Inconsistency

According to the network plots (Figure 1), we did not find any statistically inconsistent evidence of results in OS, PFS, or grade 3–5 adverse events (eTable 5).

## Discussion

To our knowledge, this is the first NMA and systematic review comparison of the efficacy and safety of currently available PARPis among cancer patients. PARPis were initially considered to inhibit PARP1/2 (catalytic inhibition) only through competitive binding to NAD catalytic sites. ^62, 63^ Recently, this type of drug has been shown to induce synthetic lethality, mainly by inhibiting PAR acylation in HRD cancers. Specifically, DNA is captured by PARP molecules (especially PARP1), which avoids further recombination with other members of the DNA repair system. Abnormal PARP1-DNA complexes induce replication fork arrest, leading to the production of DNA double-strand breaks and ultimately leading to cytotoxic effects.^3^

It should be emphasized that the main therapeutic effect of PARPis seems to be related to PAR acylation and subsequent PARP capture.^64, 65^ Different PARPis show different capture capabilities, with talazoparib > niraparib > olaparib > rucaparib > veliparib; the latter basically lacks the capture capability of PARPis.^65–67^ This is completely consistent with our OS results, but in terms of PFS, we found that the benefits of rucaparib were relatively good, which may be because the patient population of rucaparib (3 studies) all had HRD or BRCA gene mutations and were treated with a monotherapy, which is consistent with the conclusions of our subsequent subgroup analysis. Surprisingly, although iniparib has significant therapeutic effects, it has not demonstrated the typical characteristics of PARPis, including the ability to selectively kill cells with HRD or inhibit the synthesis of poly (ADP ribose) polymers in intact cells,^68, 69^ and is a poor inhibitor of PARP activity.^70^ These results raise doubts about the therapeutic efficacy of iniparib and should be used with caution in this population. Overall, PARPis showed moderate OS benefits and significant PFS benefits among all cancer patients.

Due to the complex clinical background, the treatment results of PARPis were inconsistent, and therefore we conducted a subgroup analysis.^4–10^ Patients with HRD or BRCA gene mutations were more likely to benefit from PARPis, while patients without related gene mutations only benefitted in PFS. In the former group, DNA is only repaired through non-homologous end joining and single-strand annealing, which leads to a gradual accumulation of DNA damage, chromosome instability, cell mutations, and subsequent tumor transformation.^71, 72^ During this process, PARPis inhibit PAR acylation, leading to cell death via apoptosis.^73, 74^ We also found that newly diagnosed patients using PARPis as a first-line treatment achieved a better OS, while patients with a recurrence using PARPis as a follow-up treatment achieved a better PFS. PARPis monotherapy was associated with better OS and PFS benefits than combination treatment. In addition, if there was sensitivity to platinum therapy, regardless of the sensitivity level, better PFS benefits were obtained.

We conducted a comprehensive analysis of safety based on previous studies^5^. PARPi monotherapy was more likely to lead to serious AEs. This was different from our general clinical cognition, which may be related to the dosage and duration of drug use. Different types of PARPis lead to a diversity of AEs, and our study found that iniparib and veliparib were not associated with severe AEs; olaparib was associated with an increase in mild AEs; while rucaparib, talazoparib, and niraparib were associated with a significant increase in severe AEs, which was essentially consistent with the drugs’ PAR acylation and PARP capture abilities. Specifically, talazoparib was most likely to cause anemia, veliparib had the highest association with neutropenia and leukopenia, and niraparib was associated with severe thrombocytopenia.

PARPis are prone to serious side effects during use, but their toxicity can be reduced by certain clinical measures. The first cycle of treatment is usually the peak period for most adverse events, during which close monitoring of the patient’s hematological indicators can effectively prevent serious adverse events. At the same time, the treatment window of PARPis currently in use is very wide, and there is sufficient time to adjust the drug dosage. More specifically, toxicity can be effectively controlled through dose reduction and targeted supportive care. Therefore, combining patient clinical data and taking certain clinical measures to reduce PARPis toxicity can provide personalized treatment for patients.

### Limitations

Our research has limitations. First, we did not report all levels of AEs, treatment interruptions, or patient quality of life. Second, due to the lack of mature relevant data, the optimal dosage and duration of PARPis, as well as the optimal combination therapy plan, remains an unresolved issue. Third, the follow-up time for different trials may vary, which can lead to bias in estimating the long-term efficacy of the drug. Fourth, our analysis was conducted using research-level data rather than individual patient data, which may have led to high heterogeneity in our subgroup analyses.

## Conclusions

Talazoparib may be the preferred PARPis for general cancer patients, and was associated with a better efficacy and similar toxicity trends compared to physician selected treatment. PARPis showed greater potential benefits in populations with HRD and BRCA gene mutations, new diagnoses, or sensitivity to previous platinum therapy. PARPi monotherapy showed a high efficacy but was likely to cause serious adverse reactions. It is crucial to have a comprehensive understanding of the mechanisms, indications, and safety of PARPs for the treatment of cancer patients in different clinical settings.

## Data Availability

Databases (PubMed, Cochrane, Embase, etc.) were searched from Jan 1, 1980 to Oct 20, 2023, for randomized clinical trials.

## Contribution

GQX and JWY wrote the protocol, managed the literature searches, analyzed data and wrote the draft of the manuscript. XPY, YYY, ZDC and XYC managed the literature searches and analyses. Data extraction was done by MLW, RLZ, MFY. ZKX and MFY undertook the statistical analysis. LZ, JZ and MFY modified the manuscript. All authors contributed to and have approved the final manuscript.

## Supple

**eTable 1.** PRISMA NMA Checklist of Items to Include When Reporting a Systematic Review Involving a Network Meta-Analysis.

| Section/Topic | Item # | Checklist Item | Reported on Page # |
| --- | --- | --- | --- |
| **Title** |  |  |  |
| Title | 1 | Identify the report as a systematic review incorporating a network meta-analysis (or related form of meta-analysis). | 1 |
| **Abstract** |  |  |  |
| Structured summary | 2 | Background: main objectives<br>**Methods:** data sources; study eligibility criteria, participants, and interventions; study appraisal; and synthesis methods, such as network meta-analysis.<br>**Results:** number of studies and participants identified; summary estimates with corresponding confidence/credible intervals; treatment rankings may also be discussed. Authors may choose to summarize pairwise comparisons against a chosen treatment included in their analyses for brevity.<br>**Discussion/Conclusions:** limitations; conclusions and implications of findings.<br>**Other:** primary source of funding; systematic review registration number with registry name. | 3 |
| **Introduction** |  |  |  |
| Rationale | 3 | Describe the rationale for the review in the context of what is already known, including mention of why a network meta-analysis has been conducted. | 5 |
| Objectives | 4 | Provide an explicit statement of questions being addressed, with reference to participants, interventions, comparisons, outcomes, and study design (PICOS). | 5 |
| **Methods** |  |  |  |
| Protocol and registration | 5 | Indicate whether a review protocol exists and if and where it can be accessed (e.g., Web address); and, if available, provide registration information, including registration number. | 6 |
| Eligibility criteria | 6 | Specify study characteristics (e.g., PICOS, length of follow-up) and report characteristics (e.g., years considered, language, publication status) used as criteria for eligibility, giving rationale. Clearly describe eligible treatments included in the treatment network and note whether any have been clustered or merged into the same node (with justification). | 6 |
| Information sources | 7 | Describe all information sources (e.g., databases with dates of coverage, contact with study authors to identify additional studies) in the search and date last searched. | 6 |
| Search | 8 | Present full electronic search strategy for at least one database, including any limits used, such that it could be repeated. | 6 |
| Study selection | 9 | State the process for selecting studies (i.e., screening, eligibility, included in systematic review, and, if applicable, included in the meta-analysis). | 6 |
| Data collection process | 10 | Describe method of data extraction from reports (e.g., piloted forms, independently, in duplicate) and any processes for obtaining and confirming data from investigators. | 6 |
| Data items | 11 | List and define all variables for which data were sought (e.g., PICOS, funding sources) and any assumptions and simplifications made. | 6 |
| Geometry of the network | S1 | Describe methods used to explore the geometry of the treatment network under study and potential biases related to it.<br>This should include how the evidence base has been graphically summarized for presentation, and what characteristics were compiled and used to describe the evidence base to readers. | 6 |
| Risk of bias within individual studies | 12 | Describe methods used for assessing risk of bias of individual studies (including specification of whether this was done at the study or outcome level), and how this information is to be used in any data synthesis. | 6–7 |
| Summary measures | 13 | State the principal summary measures (e.g., risk ratio, difference in means). Also describe the use of additional summary measures assessed, such as treatment rankings and surface under the cumulative ranking | 7 |
| Planned methods of analysis | 14 | Describe the methods of handling data and combining results of studies for each network meta-analysis. This should include, but not be limited to:<br>• Handling of multi-arm trials; | 7 |
|  |  | • Selection of variance structure;<br>• Selection of prior distributions in Bayesian analyses; and<br>• Assessment of model fit. |  |
| **Assessment of Inconsistency** |  | Describe the statistical methods used to evaluate the agreement of direct and indirect evidence in the treatment network(s) studied. Describe efforts taken to address its presence when found. | 7 |
| Risk of bias across studies | 15 | Specify any assessment of risk of bias that may affect the cumulative evidence (e.g., publication bias, selective reporting within studies). | 7 |
| Additional analyses | 16 | Describe methods of additional analyses if done, indicating which were pre-specified. This may include, but not be limited to, the following:<br>• Sensitivity or subgroup analyses;<br>• Meta-regression analyses;<br>• Alternative formulations of the treatment network; and<br>• Use of alternative prior distributions for Bayesian analyses (if applicable). | 7 |
| **RESULTS<sup>a</sup>** |  |  |  |
| Study selection | 17 | Give numbers of studies screened, assessed for eligibility, and included in the review, with reasons for exclusions at each | 8 |
|  |  | stage, ideally with a flow diagram. | eFigure 1 |
| Presentation of network structure |  | Provide a network graph of the included studies to enable visualization of the geometry of the treatment network. | 8<br>Figure 1 |
| Summary of network geometry |  | Provide a brief overview of characteristics of the treatment network. This may include commentary on the abundance of trials and randomized patients for the different interventions and pairwise comparisons in the network, gaps of evidence in the treatment network, and potential biases reflected by the network structure. | 8 |
| Study characteristics | 18 | For each study, present characteristics for which data were extracted (e.g., study size, PICOS, follow-up period) and provide the citations. | 8<br>eTable 3 |
| Risk of bias within studies | 19 | Present data on risk of bias of each study and, if available, any outcome level assessment. | 11<br>eFigure 2 |
| Results of individual studies | 20 | For all outcomes considered (benefits or harms), present, for each study: 1) simple summary data for each intervention group, and 2) effect estimates and confidence intervals. Modified approaches maybe needed to deal with information from larger networks. | 8–11<br>Figure 2–4<br>Table 1 |
| Synthesis of results | 21 | Present results of each meta-analysis done, including confidence/credible intervals. In larger networks, authors may focus on comparisons versus a particular comparator (e.g. placebo or standard care), with full findings presented in an appendix. League tables and forest plots maybe considered to summarize pairwise comparisons. If additional summary measures were explored (such as treatment rankings), these should also be presented. | 8–11<br>Figure 2–4<br>Table 1 |
| Exploration for inconsistency | **S5** | Describe results from investigations of inconsistency. This may include such information as measures of model fit to compare consistency and inconsistency models, P values from statistical tests, or summary of inconsistency estimates from different parts of the treatment network. | 12<br>eTable 5 |
| Risk of bias across studies | 22 | Present results of any assessment of risk of bias across studies for the evidence base being studied. | 11<br>eFigure 2 |
| Results of additional analyses | 23 | Give results of additional analyses, if done (e.g., sensitivity or subgroup analyses, meta-regression analyses, alternative network geometries studied, alternative choice of prior distributions for Bayesian analyses, and so forth). | 8–11<br>Table 1 |
| **Discussion** |  |  |  |
| Summary of evidence | 24 | Summarize the main findings, including the strength of evidence for each main outcome; consider their relevance to key groups (e.g., healthcare providers, users, and policy-makers). | 12–14 |
| Limitations | 25 | Discuss limitations at study and outcome level (e.g., risk of bias), and at review level (e.g., incomplete retrieval of identified research, reporting bias). Comment on the validity of the assumptions, such as transitivity and consistency. Comment on any concerns regarding network geometry (e.g., avoidance of certain comparisons). | 14 |
| Conclusions | 26 | Provide a general interpretation of the results in the context of other evidence, and implications for future research. | 15 |
| Funding |  |  |  |
| Funding | 27 | Describe sources of funding for the systematic review and other support (e.g., supply of data); role of funders for the systematic review. This should also include information regarding whether funding has been received from manufacturers of treatments in the network and/or whether some of the authors are content experts with professional conflicts of interest that could affect use of treatments in the network. | 15 |

PICOS = population, intervention, comparators, outcomes, study design.
ControlText in italics indicates wording specific to reporting of network meta-analyses that has been added to guidance from the PRISMA statement.
<sup>a</sup>Authors may wish to plan for use of appendices to present all relevant information in full detail for items in this section.

**eTable 2.**
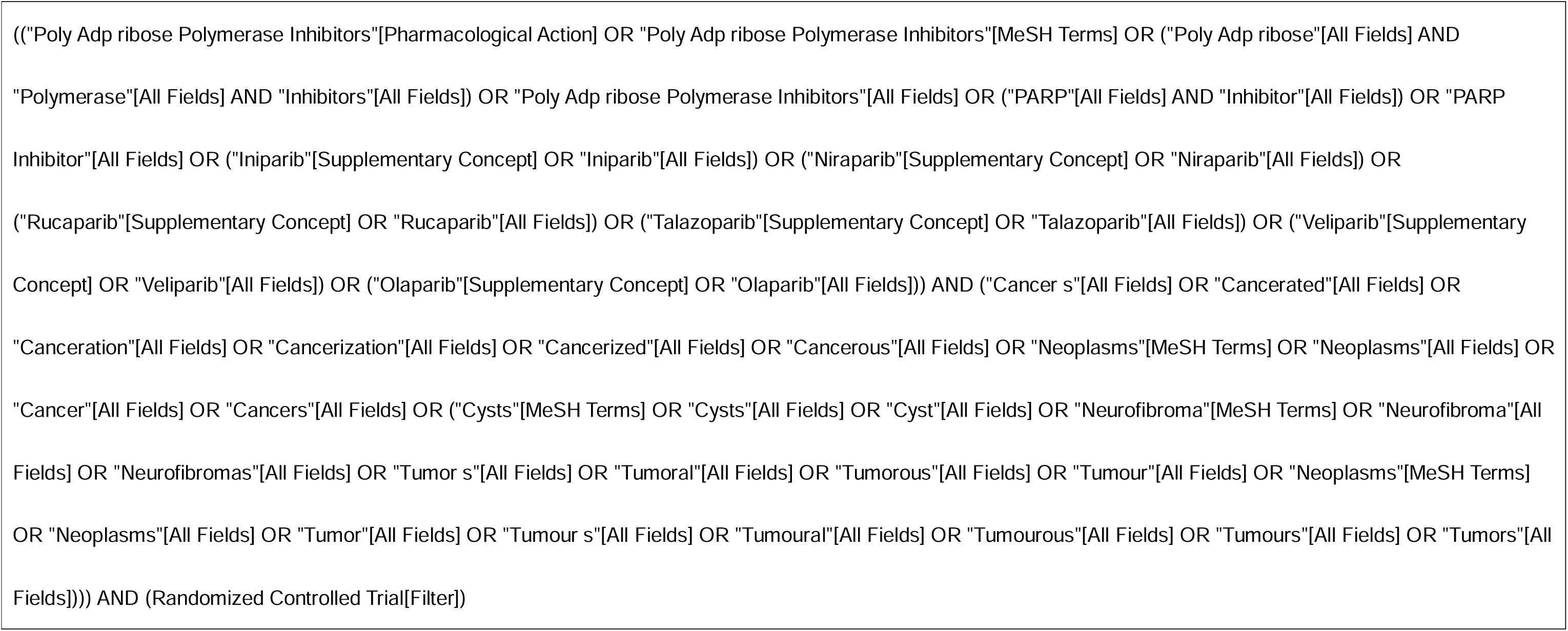
Literature Search Strategy.

**eTable 3.** Baseline Characteristics of Studies Included in the Network Meta-Analysis.

| EN | NCT | References<br>(author, y) | Country | SS | Type<br>of CA | Type of<br>MUT | ND or<br>RC | Previous<br>Platinum<br>therapy | Test<br>group | CTL | MT or<br>CB | Common<br>therapy<br>Framework | Outcomes<br>of interest | Median<br>duration of<br>follow-up, mo |
| --- | --- | --- | --- | --- | --- | --- | --- | --- | --- | --- | --- | --- | --- | --- |
| ARIEL3 | 01968213 | Oaknin et<br>al., (2021) | Spain | 564 | OC | HRD<br>BRCA MUT | RC | PR<br>CR | Rucaparib | PBO | MT | NR | PFS AE | NR |
| ARIEL4 | 02855944 | Kristeleit et<br>al., (2022) | UK | 349 | OC | BRCA1<br>BRCA2 | NR | NR | Rucaparib | TPC | MT | NR | PFS AE | 25 |
| ATHENA-MONO<br>NRGOG-3020N<br>RENGOT-ov45 | 03522246 | Monk et al.,<br>(2022) | USA | 538 | OC | HRD<br>BRCA MUT | ND | No disease<br>PR<br>CR | Rucaparib | PBO | MT | NR | PFS AE | 26.1 |
| BAYOU | 03459846 | Rosenberg<br>et al., (2022) | USA | 154 | UC | WT<br>HRD | NR | Untreated | Olaparib | PBO | CB | Imfinzi | OS PFS<br>AE | 9.8 |
| BRAVO | 01905592 | Turner et al.,<br>(2021) | UK | 215 | BC | BRCA MUT<br>BRCA1 | NR | Untreated | Niraparib | TPC | MT | NR | OS PFS<br>AE | 21.4 |
|  |  |  |  |  |  | BRCA2 |  |  |  |  |  |  |  |  |
| BrighTNess | 02032277 | Geyer et al.,<br>(2022) | USA | 476 | BC | NR | NR | NR | Veliparib | PBO | CB | PTX+CBP | OS | 54 |
| BrighTNess | 02032277 | Loibl et al.,<br>(2018) | German | 476 | BC | NR | NR | NR | Veliparib | PBO | CB | PTX+CBP | AE | 54 |
| BROCADE3 | 02163694 | Han et al.,<br>(2022) | USA | 509 | BC | BRCA MUT | NR | NR | Veliparib | PBO | CB | CBP/PTX | PFS | NR |
| BROCADE3 | 02163694 | Ayoub et al.,<br>(2021) | Canada | 509 | BC | BRCA1<br>BRCA2 | NR | NR | Veliparib | PBO | CB | CBP/PTX | OS AE | NR |
| CLIONRBGOG-ov10 | 02822157 | Vanderstich-<br>ele et al.,<br>(2022) | Belgium | 160 | OC | NR | RC | NR | Olaparib | TPC | MT | NR | OS PFS | 34 |
| ECOG-ACRIN<br>2511 | 01642251 | Owonikoko<br>et al., (2018) | USA | 128 | SCLC | NR | ND | NR | Veliparib | PBO | CB | CDDP+<br>VP-16 | OS PFS<br>AE | 18.5 |
| EMBRACA | 01945775 | Lee et al., | Korea | 431 | BC | NR | NR | NR | Talazopari | TPC | MT | NR | OS PFS | NR |
|  |  | (2021) |  |  |  |  |  |  | b |  |  |  | AE |  |
| EN | NCT | References<br>(author, y) | Country | SS | Type<br>of CA | Type of<br>MUT | ND or<br>RC | Sensitivity<br>to platinum<br>therapy | Test<br>group | CTL | MT or<br>CB | Common<br>therapy<br>Framework | Efficacy<br>outcomes<br>of interest | Median<br>duration of<br>follow-up, mo |
| ENGOT-OV16N<br>RNOVA | 01847274 | Del et al.,<br>(2019) | Spain | 553 | OC | WT<br>BRCA MUT | RC | PR<br>CR | Niraparib | PBO | MT | NR | PFS | 17.3 |
| GeparOLA | 02789332 | Fasching et<br>al., (2020 ) | German | 107 | BC | NR | NR | NR | Olaparib | TPC | CB | PTX+EPB/<br>CTX | AE | NR |
| GOAL | 01513174 | Garcia et al.,<br>(2020) | Spain | 182 | NSCL<br>C | NR | NR | NR | Olaparib | Cont<br>rol | CB | Gefitinib | OS PFS<br>AE | 12 |
| GOLD | 01924533 | Bang et<br>al.,(2017) | Korea | 525 | GC | NR | NR | NR | Olaparib | PBO | CB | PTX | OS PFS<br>AE | 11.1 |
| NORA | 03705156 | Wu et al.,<br>(2021) | China | 265 | OC | WT<br>BRCA MUT | RC | PR<br>CR | Niraparib | PBO | MT | NR | PFS AE | 15.8 |
| NRG-GY004 | 02446600 | Liu et al.,<br>(2022) | USA | 376 | OC | WT<br>BRCA MUT | RC | NR | Olaparib | TPC | MT | NR | PFS AE | 24 |
| OlympiA | 02032823 | Tutt et al.,<br>(2021) | USA | 183<br>6 | BC | NR | NR | NR | Olaparib | PBO | MT | NR | OS AE | 30 |
| OlympiAD | 02000622 | Robson et<br>al., (2023) | USA | 302 | BC | BRCA MUT | NR | NR | Olaparib | TPC | MT | NR | OS | 18.9 |
| OlympiAD | 02000622 | Im et al.,<br>(2020) | Korea | 302 | BC | NR | NR | NR | Olaparib | TPC | MT | NR | PFS AE | 19.3 |
| PAOLA-1NREN<br>GOT-ov25 | 02477644 | Ray et al.,<br>(2019) | France | 806 | OC | WT<br>HRD<br>BRCA MUT | ND | No disease<br>PR<br>CR | Olaparib | PBO | CB | Avastin | PFS AE | 22.9 |
| PIN | 01788332 | Fennell et<br>al., (2022) | UK | 70 | NSCL<br>C | NR | NR | NR | Olaparib | PBO | MT | NR | PFS | 6 |
| POLO | 02184195 | Kindler et<br>al., (2022) | USA | 154 | PAC | BRCA MUT<br>BRCA1<br>BRCA2 | NR | NR | Olaparib | PBO | MT | NR | OS PFS<br>AE | 31.3 |
| PRIMANR<br>ENGOT-OV26N | 02655016 | González<br>et al., (2019) | Spain | 733 | OC | WT<br>HRD | ND | PR<br>CR | Niraparib | PBO | MT | NR | PFS AE | 13.8 |
| RGOG-3012 |  |  |  |  |  | BRCA MUT |  |  |  |  |  |  |  |  |
| SOLO1NRGOG<br>3004 | 01844986 | Banerjee et al., (2021) | UK | 391 | OC | BRCA MUT | ND | NR | Olaparib | PBO | MT | NR | PFS AE | 60 |
| EN | NCT | References<br>(author, y) | Country | SS | Type<br>of CA | Type of<br>MUT | ND or<br>RC | Sensitivity<br>to platinum<br>therapy | Test<br>group | CTL | MT or<br>CB | Common<br>therapy<br>Framework | Efficacy<br>outcomes<br>of interest | Median<br>duration of<br>follow-up, mo |
| SOLO2NRENG<br>OT-Ov21 | 01874353 | Poveda et al., (2021) | UK | 295 | OC | BRCA MUT | RC | NR | Olaparib | PBO | MT | NR | OS AE | 65.7 |
| SOLO2NRENG<br>OT-Ov21 | 01874353 | Pujade et al., (2017) | France | 295 | OC | BRCA MUT | RC | NR | Olaparib | PBO | MT | NR | PFS | 22.2 |
| SOLTI<br>NeoPARP | 01204125 | Llombart et al., (2015) | Spain | 141 | BC | NR | NR | NR | Iniparib | CTL | CB | PTX | AE | NR |
| STOMP | ISRCTN<br>73164486 | Woll et al.,<br>(2022) | UK | 220 | SCLC | NR | NR | NR | Olaparib | PBO | MT | NR | OS PFS<br>AE | 37 |
| VELIANRGOG-3 | 02470585 | Aghajanian | USA | 757 | OC | NR | NR | NR | Veliparib | PBO | CB | CBP/PTX | AE | 28 |
| 005 |  | et al., (2021) |  |  |  |  |  |  |  |  |  |  |  |  |
| VELIANRGOG-3<br>005 | 02470585 | Coleman et<br>al., (2019 ) | USA | 757 | OC | WT<br>HRD<br>BRCA1<br>BRCA2 | ND | NR | Veliparib | PBO | CB | CBP/PTX | PFS | 28 |
| ZL-2306-005 | 03516084 | Ai et al.,<br>(2021) | China | 185 | SCLC | NR | NR | PR | Niraparib | PBO | MT | NR | OS PFS<br>AE | 7.7 |
| NR | 02289690 | Byers et al.,<br>(2021) | USA | 181 | SCLC | NR | NR | NR | Veliparib | PBO | CB | CBP+<br>VP-16 | OS PFS<br>AE | NR |
| NR | 01063517 | Bang et al.,<br>(2015) | Korea | 124 | GC | NR | NR | NR | Olaparib | PBO | CB | PTX | OS PFS<br>AE | 8.4 |
| NR | 01972217 | Clarke et al.,<br>(2018) | UK | 142 | PRC | WT<br>HRD | NR | NR | Olaparib | PBO | CB | Abiraterone | OS PFS<br>AE | 15.9 |
| NR | 02305758 | Gorbunova<br>et al., (2018) | Russia | 130 | CRC | NR | NR | NR | Veliparib | PBO | CB | FOLFIRI±<br>Avastin | OS PFS<br>AE | 23.8 |
| NR | 01506609 | Han et al., | USA | 196 | BC | BRCA MUT | NR | NR | Veliparib | PBO | CB | CBP+PTX | OS PFS | NR |
|  |  | (2018) |  |  |  |  |  |  |  |  |  |  | AE |  |
| NR | 00753545 | Ledermann<br>et al.,(2014) | UK | 265 | OC | WT<br>BRCA MUT | RC | NR | Olaparib | PBO | MT | NR | OS PFS<br>AE | 37.3 |
| NR | 00804908 | Middleton et<br>al.,(2015) | UK | 346 | Mel | NR | NR | NR | Veliparib | PBO | CB | TMZ | OS PFS<br>AE | NR |
| EN | NCT | References<br>(author, y) | Country | SS | Type<br>of CA | Type of<br>MUT | ND or<br>RC | Sensitivity<br>to platinum<br>therapy | Test<br>group | CTL | MT or<br>CB | Common<br>therapy<br>Framework | Efficacy<br>outcomes<br>of interest | Median<br>duration of<br>follow-up, mo |
| NR | 01086254 | Novello et<br>al., (2014) | Italy | 119 | NSCL<br>C | NR | NR | NR | Iniparib | CTL | CB | GEM+<br>CDDP | OS PFS<br>AE | NR |
| NR | 00938652 | O'Shaughne<br>ssy et al.,<br>(2014) | USA | 519 | BC | NR | NR | NR | Iniparib | CTL | CB | GEM+CBP | OS PFS<br>AE | NR |
| NR | 00540358 | O'Shaughne<br>ssy et<br>al.,(2011 ) | USA | 123 | BC | NR | NR | NR | Iniparib | CTL | CB | GEM+CBP | OS PFS<br>AE | NR |
| NR | 01081951 | Oza et al.,<br>(2014) | Canada | 162 | OC | BRCA MUT | RC | NR | Olaparib | CTL | CB | CBP+PTX | OS PFS<br>AE | 44 |
| NR | 01638546 | Pietanza et<br>al., (2018) | USA | 104 | SCLC | NR | NR | NR | Veliparib | PBO | CB | TMZ | OS PFS<br>AE | NR |
| NR | 02106546 | Ramalingam<br>et al., (2021) | USA | 970 | NSCL<br>C | NR | NR | NR | Veliparib | PBO | CB | CBP/PTX | OS PFS<br>AE | 49.8 |
| NR | 01560104 | Ramalingam<br>et al.,(2017) | USA | 158 | NSCL<br>C | NR | NR | NR | Veliparib | PBO | CB | CBP+PTX | OS PFS<br>AE | NR |

AE, Adverse event; BC, Breast Cancer; CB, Combination; CA, Cancer; CBP, Carboplatin ; CDDP, cisplatin; CR, Complete Response; CRC, Colorectal cancer; CTL, Control Group; CTX, Cyclophosphamide; EN,
Experiment name; EPB, Epirubicin; FOLFIRI, Folinic acid+Fluorouracil +Irinotecan; GC, Gastric Cancer; GEM, Gemcitabine; HRD, Homologous Recombination Deficiency; Mel, Melanoma; MT, Monotherapy; MUT,
mutation; NCT, National Clinical Trial; ND, New diagnosis; NR, not reported; NSCLC, Non-Small Cell Lung Cancer; OC, Ovarian Cancer; OS, Overall survival; PAC, Pancreatic Cancer; PBO, Placebo; PFS,
Progression-free survival; PR, Partial Response; PRC, Prostate cancer; PTX, Paclitaxel; RC, Recurrence; SCLC, Small Cell Lung Cancer; SS, Sample size; TPC, Treatment of physician's choice; TMZ, Temozolomide;
UC, Urothelial carcinoma; VP-16, Etoposide; WT, Wild Type; y, year; Control represents that no longer receive other drugs except under the same treatment medication as the experimental group.

**eTable 4.** Ranking Profiles in the Bayesian Network Meta-Analysis.

| **Overall Survival** | **Rank Probability (%)** |  |  |  |  |  |  |  |  |
| --- | --- | --- | --- | --- | --- | --- | --- | --- | --- |
| Therapeutic drugs | Rank 1<sup>st</sup> | Rank 2<sup>nd</sup> | Rank 3<sup>rd</sup> | Rank 4<sup>th</sup> | Rank 5<sup>th</sup> | Rank 6<sup>th</sup> | Rank 7<sup>th</sup> | Rank 8<sup>th</sup> | Rank 9<sup>th</sup> |
| Olaparib | 0.02 | 0.14 | 0.30 | 0.28 | 0.18 | 0.07 | 0.00 | 0.00 | NA |
| Veliparib | 0.00 | 0.01 | 0.04 | 0.11 | 0.20 | 0.28 | 0.35 | 0.01 | NA |
| Talazoparib | 0.23 | 0.26 | 0.17 | 0.12 | 0.10 | 0.06 | 0.04 | 0.03 | NA |
| Iniparib | 0.69 | 0.17 | 0.06 | 0.04 | 0.02 | 0.01 | 0.01 | 0.00 | NA |
| Niraparib | 0.06 | 0.13 | 0.16 | 0.16 | 0.16 | 0.15 | 0.10 | 0.08 | NA |
| TPC | 0.00 | 0.04 | 0.09 | 0.13 | 0.19 | 0.21 | 0.17 | 0.16 | NA |
| Placebo | 0.00 | 0.00 | 0.00 | 0.00 | 0.02 | 0.09 | 0.24 | 0.64 | NA |
| Control | 0.00 | 0.25 | 0.19 | 0.15 | 0.13 | 0.12 | 0.08 | 0.08 | NA |
| **Progression-free Survival** | **Rank Probability (%)** |  |  |  |  |  |  |  |  |
| Therapeutic drugs | Rank 1<sup>st</sup> | Rank 2<sup>nd</sup> | Rank 3<sup>rd</sup> | Rank 4<sup>th</sup> | Rank 5<sup>th</sup> | Rank 6<sup>th</sup> | Rank 7<sup>th</sup> | Rank 8<sup>th</sup> | Rank 9<sup>th</sup> |
| Olaparib | 0.00 | 0.00 | 0.00 | 0.02 | 0.20 | 0.54 | 0.25 | 0.00 | 0.00 |
| Veliparib | 0.00 | 0.00 | 0.00 | 0.00 | 0.00 | 0.00 | 0.08 | 0.91 | 0.00 |
| Talazoparib | 0.93 | 0.06 | 0.01 | 0.00 | 0.00 | 0.00 | 0.00 | 0.00 | 0.00 |
| Iniparib | 0.01 | 0.09 | 0.40 | 0.28 | 0.13 | 0.09 | 0.00 | 0.00 | 0.00 |
| Niraparib | 0.00 | 0.01 | 0.45 | 0.44 | 0.09 | 0.01 | 0.00 | 0.00 | 0.00 |
| TPC | 0.00 | 0.00 | 0.04 | 0.22 | 0.45 | 0.20 | 0.09 | 0.00 | 0.00 |
| Placebo | 0.00 | 0.00 | 0.00 | 0.00 | 0.00 | 0.00 | 0.00 | 0.00 | 1.00 |
| Control | 0.00 | 0.00 | 0.00 | 0.04 | 0.14 | 0.16 | 0.58 | 0.09 | 0.00 |
| Rucaparib | 0.06 | 0.83 | 0.10 | 0.00 | 0.00 | 0.00 | 0.00 | 0.00 | 0.00 |
| **Grade 3–5 Adverse Events** | **Rank Probability (%)** |  |  |  |  |  |  |  |  |
| Therapeutic drugs | Rank 1<sup>st</sup> | Rank 2<sup>nd</sup> | Rank 3<sup>rd</sup> | Rank 4<sup>th</sup> | Rank 5<sup>th</sup> | Rank 6<sup>th</sup> | Rank 7<sup>th</sup> | Rank 8<sup>th</sup> | Rank 9<sup>th</sup> |
| Olaparib | 0.00 | 0.00 | 0.00 | 0.02 | 0.32 | 0.33 | 0.28 | 0.05 | 0.00 |
| Veliparib | 0.00 | 0.00 | 0.00 | 0.01 | 0.08 | 0.19 | 0.21 | 0.47 | 0.05 |
| Talazoparib | 0.23 | 0.24 | 0.22 | 0.20 | 0.06 | 0.03 | 0.02 | 0.01 | 0.00 |
| Iniparib | 0.01 | 0.03 | 0.06 | 0.11 | 0.35 | 0.16 | 0.14 | 0.09 | 0.06 |
| Niraparib | 0.09 | 0.32 | 0.24 | 0.28 | 0.05 | 0.01 | 0.00 | 0.00 | 0.00 |
| TPC | 0.02 | 0.16 | 0.40 | 0.36 | 0.05 | 0.01 | 0.00 | 0.00 | 0.00 |
| Placebo | 0.00 | 0.00 | 0.00 | 0.00 | 0.00 | 0.01 | 0.07 | 0.15 | 0.77 |
| Control | 0.00 | 0.00 | 0.01 | 0.02 | 0.09 | 0.26 | 0.28 | 0.23 | 0.12 |
| Rucaparib | 0.65 | 0.26 | 0.07 | 0.02 | 0.00 | 0.00 | 0.00 | 0.00 | 0.00 |

Each box represents the possibility of the row-defining treatments versus the column-definition ranking. The possibility of each box is related to the color scale, with green indicating low, yellow showing moderate, and red representing high. TPC, Treatment of physician's choice; Control represents that no longer receive other drugs except under the same treatment medication as the experimental group.

**eTable 5.** Inconsistency Analysis of the Network Meta-Analysis Results.

|  |  |  |  |  |  |
| --- | --- | --- | --- | --- | --- |
| **Overall survival** |  |  |  |  |  |
| Loop | IF | seIF | z_value | p_value | CI |
| Niraparib-Olaparib-TPC-Placebo | 0.121 | 0.464 | 0.261 | 0.794 | 0.00-1.03 |
| **Progression-free survival** |  |  |  |  |  |
| Loop | IF | seIF | z_value | p_value | CI |
| Niraparib-Rucaparib-TPC-Placebo | 0.155 | 0.345 | 0.449 | 0.654 | 0.00-0.83 |
| Olaparib-Rucaparib-TPC-Placebo | 0.126 | 0.371 | 0.338 | 0.735 | 0.00-0.85 |
| Niraparib-Olaparib-TPC-Placebo | 0.029 | 0.295 | 0.099 | 0.921 | 0.00-0.61 |
| **Grade 3–5 adverse events** |  |  |  |  |  |
| Loop | IF | seIF | z_value | p_value | CI |
| Olaparib-Rucaparib-TPC-Placebo | 0.472 | 0.648 | 0.729 | 0.466 | 0.00-1.74 |

CI, confidence interval; IF, Inconsistency factor; TPC, Treatment of physician's choice; **seIF**, Standard Error Inconsistency factor;

**eFigure 1.**
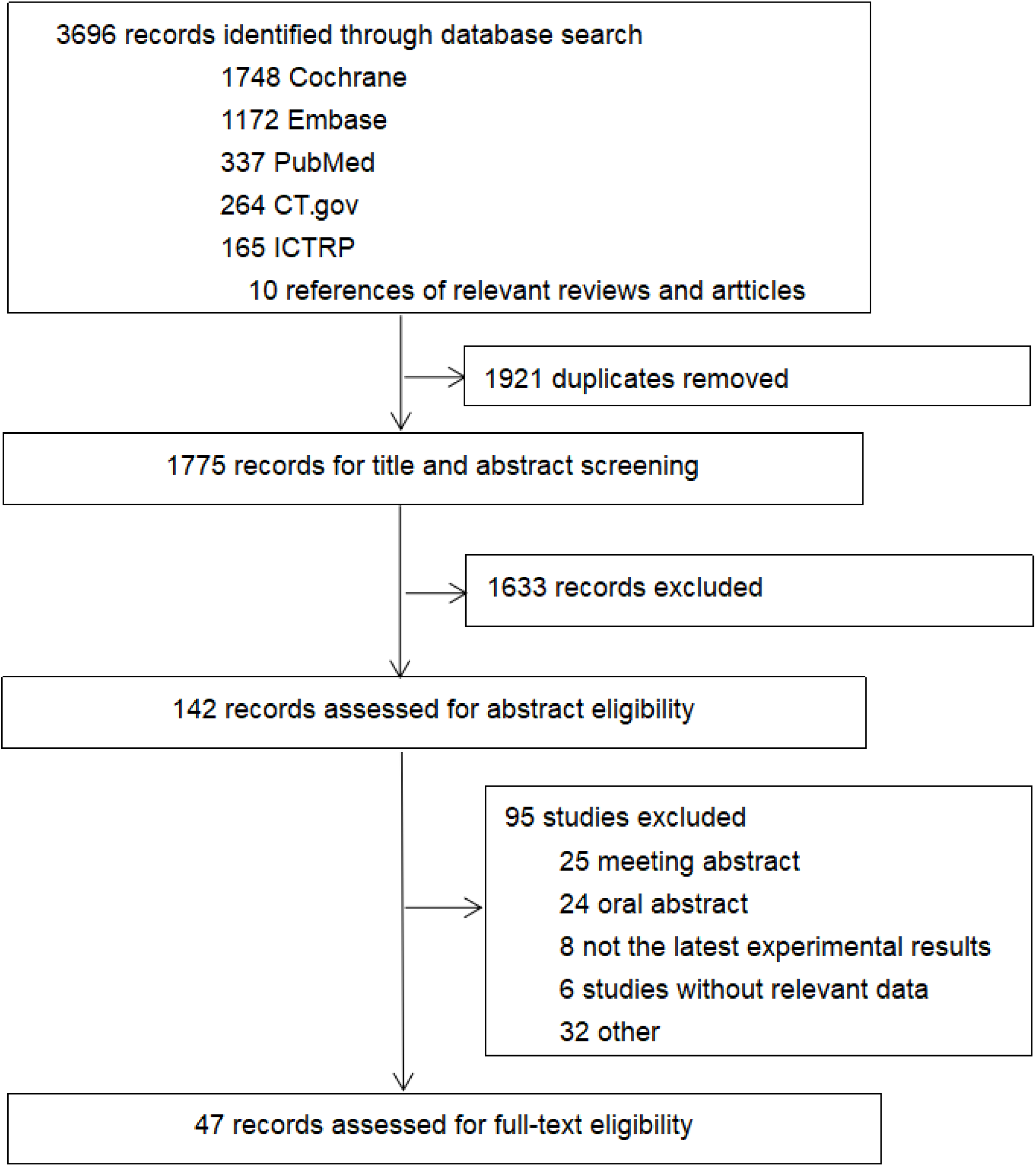
Literature search and selection.

**eFigure 2.**
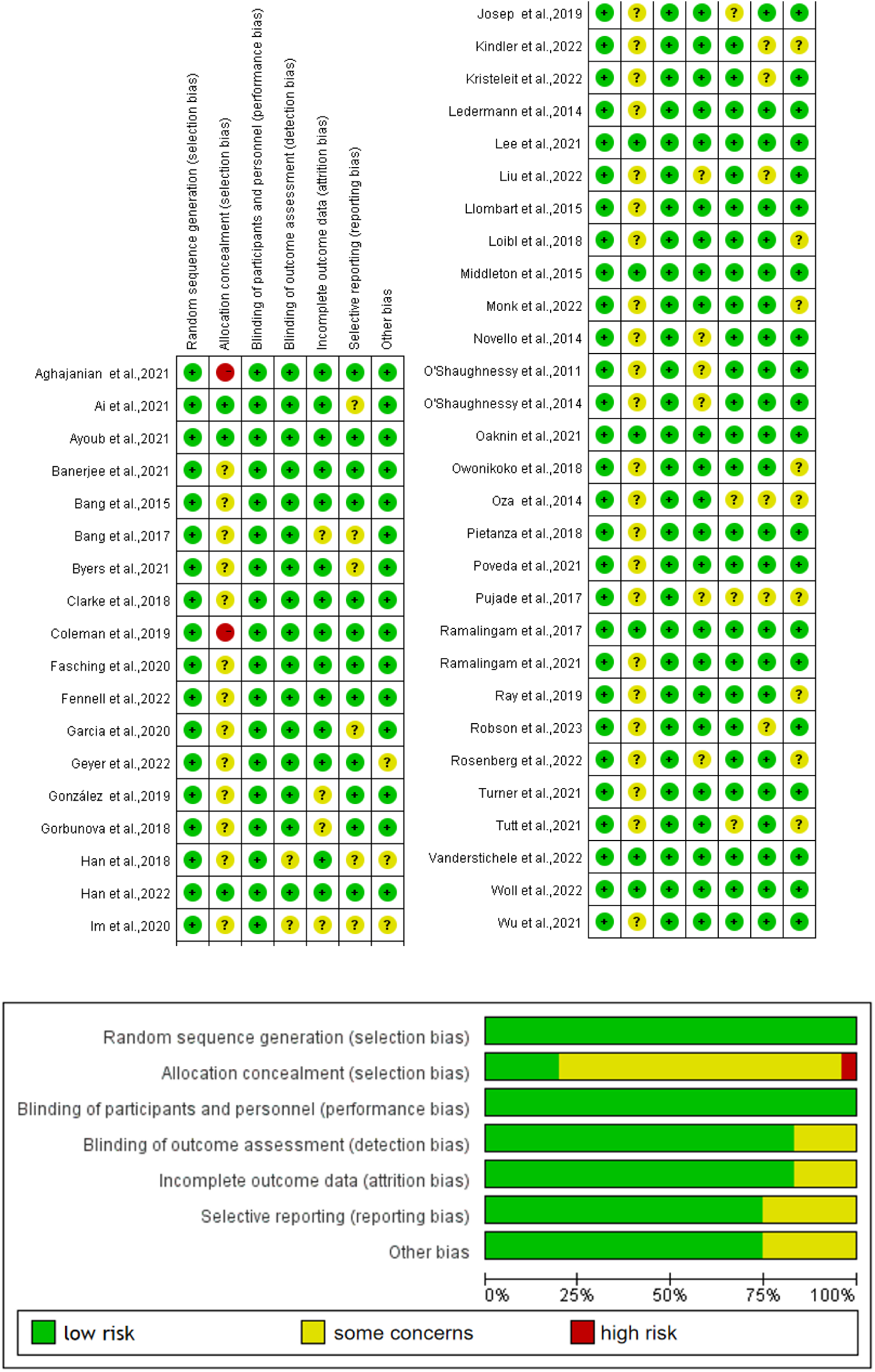
Summary of quality assessments using the Cochrane Risk of Bias Tool 2.0. Studies were classified into one of three categories: low, some concerns, or high risk.

## Declarations

### Ethical Approval and Availability of data and materials

The above content is not applicable.

## References

1. Negrini S, Gorgoulis VG, Halazonetis TD. Genomic instability--an evolving hallmark of cancer. Nat Rev Mol Cell Biol. 2010;11:220–228. doi:10.1038/nrm2858

2. Institute NC. Genetics of Breast and Gynecologic Cancers (PDQ(R)): Health Professional Version. 2002.

3. Murai J, Huang SY, Das BB, et al. Trapping of PARP1 and PARP2 by Clinical PARP Inhibitors. CANCER RES. 2012;72:5588–5599. doi:10.1158/0008-5472.CAN-12-2753

4. Ruscito I, Bellati F, Ray-Coquard I, et al. Incorporating Parp-inhibitors in Primary and Recurrent Ovarian Cancer: A Meta-analysis of 12 phase II/III randomized controlled trials. CANCER TREAT REV. 2020;87:102040. doi:10.1016/j.ctrv.2020.102040

5. Yang Y, Du N, Xie L, et al. The efficacy and safety of the addition of poly ADP-ribose polymerase (PARP) inhibitors to therapy for ovarian cancer: a systematic review and meta-analysis. WORLD J SURG ONCOL. 2020;18:151. doi:10.1186/s12957-020-01931-7

6. Shao F, Liu J, Duan Y, et al. Efficacy and safety of PARP inhibitors as the maintenance therapy in ovarian cancer: a meta-analysis of nine randomized controlled trials. Biosci Rep. 2020;40. doi:10.1042/BSR20192226

7. Lin Q, Liu W, Xu S, et al. PARP inhibitors as maintenance therapy in newly diagnosed advanced ovarian cancer: a meta-analysis. BJOG. 2021;128:485–493. doi:10.1111/1471-0528.16411

8. Tomao F, Bardhi E, Di Pinto A, et al. Parp inhibitors as maintenance treatment in platinum sensitive recurrent ovarian cancer: An updated meta-analysis of randomized clinical trials according to BRCA mutational status. CANCER TREAT REV. 2019;80:101909. doi:10.1016/j.ctrv.2019.101909

9. Sun X, Xu S, Li Y, Lv X, Wei M, He M. Efficacy and safety of PARP inhibitors in the treatment of BRCA-mutated breast cancer: an updated systematic review and meta-analysis of randomized controlled trials. Expert Rev Clin Pharmacol. 2023;16:245–256. doi:10.1080/17512433.2023.2188193

10. Sun X, Wang X, Zhang J, et al. Efficacy and safety of PARP inhibitors in patients with BRCA-mutated advanced breast cancer: A meta-analysis and systematic review. BREAST. 2021;60:26–34. doi:10.1016/j.breast.2021.08.009

11. Gulia S, Kannan S, Ghosh J, Rath S, Maheshwari A, Gupta S. Maintenance therapy with a poly(ADP-ribose) polymerase inhibitor in patients with newly diagnosed advanced epithelial ovarian cancer: individual patient data and trial-level meta-analysis. ESMO Open. 2022;7:100558. doi:10.1016/j.esmoop.2022.100558

12. Hutton B, Salanti G, Caldwell DM, et al. The PRISMA extension statement for reporting of systematic reviews incorporating network meta-analyses of health care interventions: checklist and explanations. ANN INTERN MED. 2015;162:777–784. doi:10.7326/M14-2385

13. AD HJ. Assessing risk of bias in included studies. John Wiley & Sons. 2008.

14. Higgins JP, Thompson SG, Deeks JJ, Altman DG. Measuring inconsistency in meta-analyses. BMJ. 2003;327:557–560. doi:10.1136/bmj.327.7414.557

15. Oaknin A, Oza AM, Lorusso D, et al. Maintenance treatment with rucaparib for recurrent ovarian carcinoma in ARIEL3, a randomized phase 3 trial: The effects of best response to last platinum-based regimen and disease at baseline on efficacy and safety. CANCER MED-US. 2021;10:7162–7173. doi:10.1002/cam4.4260

16. Kristeleit R, Lisyanskaya A, Fedenko A, et al. Rucaparib versus standard-of-care chemotherapy in patients with relapsed ovarian cancer and a deleterious BRCA1 or BRCA2 mutation (ARIEL4): an international, open-label, randomised, phase 3 trial. The Lancet Oncology. 2022;23:465–478. doi:10.1016/S1470-2045(22)00122-X

17. Monk BJ, Parkinson C, Lim MC, et al. A Randomized, Phase III Trial to Evaluate Rucaparib Monotherapy as Maintenance Treatment in Patients With Newly Diagnosed Ovarian Cancer (ATHENA –MONO/GOG-3020/ENGOT-ov45). J CLIN ONCOL. 2022;40:3952–3964. doi:10.1200/JCO.22.01003

18. Rosenberg JE, Park SH, Kozlov V, et al. Durvalumab Plus Olaparib in Previously Untreated, Platinum-Ineligible Patients With Metastatic Urothelial Carcinoma: A Multicenter, Randomized, Phase II Trial (BAYOU). J CLIN ONCOL. 2022;41:43–53. doi:10.1200/JCO.22.00205

19. Turner NC, Balmaña J, Poncet C, et al. Niraparib for Advanced Breast Cancer with Germline BRCA1 and BRCA2 Mutations: the EORTC 1307-BCG/BIG5–13/TESARO PR-30–50–10-C BRAVO Study. CLIN CANCER RES. 2021;27:5482–5491. doi:10.1158/1078-0432.CCR-21-0310

20. Geyer CE, Sikov WM, Huober J, et al. Long-term efficacy and safety of addition of carboplatin with or without veliparib to standard neoadjuvant chemotherapy in triple-negative breast cancer: 4-year follow-up data from BrighTNess, a randomized phase III trial. ANN ONCOL. 2022;33:384–394. doi:10.1016/j.annonc.2022.01.009

21. Loibl S, O’Shaughnessy J, Untch M, et al. Addition of the PARP inhibitor veliparib plus carboplatin or carboplatin alone to standard neoadjuvant chemotherapy in triple-negative breast cancer (BrighTNess): a randomised, phase 3 trial. The Lancet Oncology. 2018;19:497–509. doi:10.1016/S1470-2045(18)30111-6

22. Han HS, Arun BK, Kaufman B, et al. Veliparib monotherapy following carboplatin/paclitaxel plus veliparib combination therapy in patients with germline BRCA-associated advanced breast cancer: results of exploratory analyses from the phase III BROCADE3 trial. ANN ONCOL. 2022;33:299–309. doi:10.1016/j.annonc.2021.11.018

23. Ayoub J, Wildiers H, Friedlander M, et al. Safety and efficacy of veliparib plus carboplatin/paclitaxel in patients with HER2-negative metastatic or locally advanced breast cancer: subgroup analyses by germline BRCA1/2 mutations and hormone receptor status from the phase-3 BROCADE3 trial. THER ADV MED ONCOL. 2021;13:17495663. doi:10.1177/17588359211059601

24. Vanderstichele A, Loverix L, Busschaert P, et al. Randomized CLIO/BGOG-ov10 trial of olaparib monotherapy versus physician’s choice chemotherapy in relapsed ovarian cancer. GYNECOL ONCOL. 2022;165:14–22. doi:10.1016/j.ygyno.2022.01.034

25. Owonikoko TK, Dahlberg SE, Sica GL, et al. Randomized Phase II Trial of Cisplatin and Etoposide in Combination With Veliparib or Placebo for Extensive-Stage Small-Cell Lung Cancer: ECOG-ACRIN 2511 Study. J CLIN ONCOL. 2018;37:222–229. doi:10.1200/JCO.18.00264

26. Lee K, Sohn J, Goodwin A, et al. Talazoparib Versus Chemotherapy in Patients with HER2-negative Advanced Breast Cancer and a Germline BRCA1/2 Mutation Enrolled in Asian Countries: Exploratory Subgroup Analysis of the Phase III EMBRACA Trial. CANCER RES TREAT. 2021;53:1084–1095. doi:10.4143/crt.2020.1381

27. Del CJ, Matulonis UA, Malander S, et al. Niraparib Maintenance Therapy in Patients With Recurrent Ovarian Cancer After a Partial Response to the Last Platinum-Based Chemotherapy in the ENGOT-OV16/NOVA Trial. J CLIN ONCOL. 2019;37:2968–2973. doi:10.1200/JCO.18.02238

28. Fasching PA, Link T, Hauke J, et al. Neoadjuvant paclitaxel/olaparib in comparison to paclitaxel/carboplatinum in patients with HER2-negative breast cancer and homologous recombination deficiency (GeparOLA study). 2020;32:47–49. doi:10.1016/j.annonc.2020.10.471

29. Garcia-Campelo R, Arrieta O, Massuti B, et al. Combination of gefitinib and olaparib versus gefitinib alone in EGFR mutant non-small-cell lung cancer (NSCLC): A multicenter, randomized phase II study (GOAL). LUNG CANCER. 2020;150:62–69. doi:10.1016/j.lungcan.2020.09.018

30. Bang Y, Xu R, Chin K, et al. Olaparib in combination with paclitaxel in patients with advanced gastric cancer who have progressed following first-line therapy (GOLD): a double-blind, randomised, placebo-controlled, phase 3 trial. The Lancet Oncology. 2017;18:1637–1651. doi:10.1016/S1470-2045(17)30682-4

31. Wu XH, Zhu JQ, Yin RT, et al. Niraparib maintenance therapy in patients with platinum-sensitive recurrent ovarian cancer using an individualized starting dose (NORA): a randomized, double-blind, placebo-controlled phase III trial. ANN ONCOL. 2021;32:512–521. doi:10.1016/j.annonc.2020.12.018

32. Liu JF, Brady MF, Matulonis UA, et al. Olaparib With or Without Cediranib Versus Platinum-Based Chemotherapy in Recurrent Platinum-Sensitive Ovarian Cancer (NRG-GY004): A Randomized, Open-Label, Phase III Trial. J CLIN ONCOL. 2022;40:2138–2147. doi:10.1200/JCO.21.02011

33. Tutt ANJ, Garber JE, Kaufman B, et al. Adjuvant Olaparib for Patients withBRCA1 - or BRCA2-Mutated Breast Cancer. NEW ENGL J MED. 2021;384:2394–2405. doi:10.1056/NEJMoa2105215

34. Robson ME, Im SA, Senkus E, et al. OlympiAD extended follow-up for overall survival and safety: Olaparib versus chemotherapy treatment of physician’s choice in patients with a germline BRCA mutation and HER2-negative metastatic breast cancer. EUR J CANCER. 2023;184:39–47. doi:10.1016/j.ejca.2023.01.031

35. Im S, Xu B, Li W, et al. Olaparib monotherapy for Asian patients with a germline BRCA mutation and HER2-negative metastatic breast cancer: OlympiAD randomized trial subgroup analysis. SCI REP-UK. 2020;10:8753. doi:10.1038/s41598-020-63033-4

36. Ray-Coquard I, Pautier P, Pignata S, et al. Olaparib plus Bevacizumab as First-Line Maintenance in Ovarian Cancer. N Engl J Med. 2019;381:2416–2428. doi:10.1056/NEJMoa1911361

37. Fennell DA, Porter C, Lester J, et al. Olaparib maintenance versus placebo monotherapy in patients with advanced non-small cell lung cancer (PIN): A multicentre, randomised, controlled, phase 2 trial. eClinicalMedicine. 2022;52:101595. doi:10.1016/j.eclinm.2022.101595

38. Kindler HL, Hammel P, Reni M, et al. Overall Survival Results From the POLO Trial: A Phase III Study of Active Maintenance Olaparib Versus Placebo for Germline BRCA-Mutated Metastatic Pancreatic Cancer. J CLIN ONCOL. 2022;40:3929–3939. doi:10.1200/JCO.21.01604

39. González-Martín A, Pothuri B, Vergote I, et al. Niraparib in Patients with Newly Diagnosed Advanced Ovarian Cancer. NEW ENGL J MED. 2019;381:2391–2402. doi:10.1056/NEJMoa1910962

40. Banerjee S, Moore KN, Colombo N, et al. Maintenance olaparib for patients with newly diagnosed advanced ovarian cancer and a BRCA mutation (SOLO1/GOG 3004): 5-year follow-up of a randomised, double-blind, placebo-controlled, phase 3 trial. The Lancet Oncology. 2021;22:1721–1731. doi:10.1016/S1470-2045(21)00531-3

41. Poveda A, Floquet A, Ledermann JA, et al. Olaparib tablets as maintenance therapy in patients with platinum-sensitive relapsed ovarian cancer and a BRCA1/2 mutation (SOLO2/ENGOT-Ov21): a final analysis of a double-blind, randomised, placebo-controlled, phase 3 trial. The Lancet Oncology. 2021;22:620–631. doi:10.1016/S1470-2045(21)00073-5

42. Pujade-Lauraine E, Ledermann JA, Selle F, et al. Olaparib tablets as maintenance therapy in patients with platinum-sensitive, relapsed ovarian cancer and a BRCA1/2 mutation (SOLO2/ENGOT-Ov21): a double-blind, randomised, placebo-controlled, phase 3 trial. The Lancet Oncology. 2017;18:1274–1284. doi:10.1016/S1470-2045(17)30469-2

43. Llombart-Cussac A, Bermejo B, Villanueva C, et al. SOLTI NeoPARP: a phase II randomized study of two schedules of iniparib plus paclitaxel versus paclitaxel alone as neoadjuvant therapy in patients with triple-negative breast cancer. BREAST CANCER RES TR. 2015;154:351–357. doi:10.1007/s10549-015-3616-8

44. Woll P, Gaunt P, Danson S, et al. Olaparib as maintenance treatment in patients with chemosensitive small cell lung cancer (STOMP): A randomised, double-blind, placebo-controlled phase II trial. LUNG CANCER. 2022;171:26–33. doi:10.1016/j.lungcan.2022.07.007

45. Aghajanian C, Swisher EM, Okamoto A, et al. Impact of veliparib, paclitaxel dosing regimen, and germline BRCA status on the primary treatment of serous ovarian cancer – an ancillary data analysis of the VELIA trial. GYNECOL ONCOL. 2021;164:278–287. doi:10.1016/j.ygyno.2021.12.012

46. Coleman RL, Fleming GF, Brady MF, et al. Veliparib with First-Line Chemotherapy and as Maintenance Therapy in Ovarian Cancer. N Engl J Med. 2019;381:2403–2415. doi:10.1056/NEJMoa1909707

47. Ai X, Pan Y, Shi J, et al. Efficacy and Safety of Niraparib as Maintenance Treatment in Patients With Extensive-Stage SCLC After First-Line Chemotherapy: A Randomized, Double-Blind, Phase 3 Study. J THORAC ONCOL. 2021;16:1403–1414. doi:10.1016/j.jtho.2021.04.001

48. Byers LA, Bentsion D, Gans S, et al. Veliparib in Combination with Carboplatin and Etoposide in Patients with Treatment-Naïve Extensive-Stage Small Cell Lung Cancer: A Phase 2 Randomized Study. CLIN CANCER RES. 2021;27:3884–3895. doi:10.1158/1078-0432.CCR-20-4259

49. Bang Y, Im S, Lee K, et al. Randomized, Double-Blind Phase II Trial With Prospective Classification by ATM Protein Level to Evaluate the Efficacy and Tolerability of Olaparib Plus Paclitaxel in Patients With Recurrent or Metastatic Gastric Cancer. J CLIN ONCOL. 2015;33:3858–3865. doi:10.1200/JCO.2014.60.0320

50. Clarke N, Wiechno PJ, Alekseev B, et al. Olaparib combined with abiraterone in patients (pts) with metastatic castration-resistant prostate cancer (mCRPC): A randomized phase II trial. J CLIN ONCOL. 2018;36:5003. doi:10.1200/JCO.2018.36.15_suppl.5003

51. Gorbunova V, Beck JT, Hofheinz R, et al. A phase 2 randomised study of veliparib plus FOLFIRI±bevacizumab versus placebo plus FOLFIRI±bevacizumab in metastatic colorectal cancer. BRIT J CANCER. 2018;120:183–189. doi:10.1038/s41416-018-0343-z

52. Han HS, Diéras V, Robson M, et al. Veliparib with temozolomide or carboplatin/paclitaxel versus placebo with carboplatin/paclitaxel in patients with BRCA1/2 locally recurrent/metastatic breast cancer: randomized phase II study. ANN ONCOL. 2018;29:154–161. doi:10.1093/annonc/mdx505

53. Ledermann J, Harter P, Gourley C, et al. Olaparib maintenance therapy in patients with platinum-sensitive relapsed serous ovarian cancer: a preplanned retrospective analysis of outcomes by BRCA status in a randomised phase 2 trial. The Lancet Oncology. 2014;15:852–861. doi:10.1016/S1470-2045(14)70228-1

54. Middleton MR, Friedlander P, Hamid O, et al. Randomized phase II study evaluating veliparib (ABT-888) with temozolomide in patients with metastatic melanoma. ANN ONCOL. 2015;26:2173–2179. doi:10.1093/annonc/mdv308

55. Novello S, Besse B, Felip E, et al. A phase II randomized study evaluating the addition of iniparib to gemcitabine plus cisplatin as first-line therapy for metastatic non-small-cell lung cancer. ANN ONCOL. 2014;25:2156–2162. doi:10.1093/annonc/mdu384

56. O’Shaughnessy J, Schwartzberg L, Danso MA, et al. Phase III Study of Iniparib Plus Gemcitabine and Carboplatin Versus Gemcitabine and Carboplatin in Patients With Metastatic Triple-Negative Breast Cancer. J CLIN ONCOL. 2014;32:3840–3847. doi:10.1200/JCO.2014.55.2984

57. O’Shaughnessy J, Osborne C, Pippen JE, et al. Iniparib plus Chemotherapy in Metastatic Triple-Negative Breast Cancer. NEW ENGL J MED. 2011;364:205–214. doi:10.1056/NEJMoa1011418

58. Oza AM, Cibula D, Benzaquen AO, et al. Olaparib combined with chemotherapy for recurrent platinum-sensitive ovarian cancer: a randomised phase 2 trial. The Lancet Oncology. 2014;16:87–97. doi:10.1016/S1470-2045(14)71135-0

59. Pietanza MC, Waqar SN, Krug LM, et al. Randomized, Double-Blind, Phase II Study of Temozolomide in Combination With Either Veliparib or Placebo in Patients With Relapsed-Sensitive or Refractory Small-Cell Lung Cancer. J CLIN ONCOL. 2018;36:2386–2394. doi:10.1200/JCO.2018.77.7672

60. Ramalingam SS, Novello S, Guclu SZ, et al. Veliparib in Combination With Platinum-Based Chemotherapy for First-Line Treatment of Advanced Squamous Cell Lung Cancer: A Randomized, Multicenter Phase III Study. J CLIN ONCOL. 2021;39:3633–3644. doi:10.1200/JCO.20.03318

61. Ramalingam SS, Blais N, Mazieres J, et al. Randomized, Placebo-Controlled, Phase II Study of Veliparib in Combination with Carboplatin and Paclitaxel for Advanced/Metastatic Non–Small Cell Lung Cancer. CLIN CANCER RES. 2017;23:1937–1944. doi:10.1158/1078-0432.CCR-15-3069

62. Farmer H, McCabe N, Lord CJ, et al. Targeting the DNA repair defect in BRCA mutant cells as a therapeutic strategy. NATURE. 2005;434:917–921. doi:10.1038/nature03445

63. McCabe N, Turner NC, Lord CJ, et al. Deficiency in the repair of DNA damage by homologous recombination and sensitivity to poly(ADP-ribose) polymerase inhibition. CANCER RES. 2006;66:8109–8115. doi:10.1158/0008-5472.CAN-06-0140

64. Lord CJ, Ashworth A. PARP inhibitors: Synthetic lethality in the clinic. SCIENCE. 2017;355:1152–1158. doi:10.1126/science.aam7344

65. Murai J, Huang SY, Das BB, et al. Trapping of PARP1 and PARP2 by Clinical PARP Inhibitors. CANCER RES. 2012;72:5588–5599. doi:10.1158/0008-5472.CAN-12-2753

66. Murai J, Huang SY, Renaud A, et al. Stereospecific PARP trapping by BMN 673 and comparison with olaparib and rucaparib. MOL CANCER THER. 2014;13:433–443. doi:10.1158/1535-7163.MCT-13-0803

67. Pommier Y, O’Connor MJ, de Bono J. Laying a trap to kill cancer cells: PARP inhibitors and their mechanisms of action. SCI TRANSL MED. 2016;8:317p–362p. doi:10.1126/scitranslmed.aaf9246

68. Patel AG, De Lorenzo SB, Flatten KS, Poirier GG, Kaufmann SH. Failure of iniparib to inhibit poly(ADP-Ribose) polymerase in vitro. CLIN CANCER RES. 2012;18:1655–1662. doi:10.1158/1078-0432.CCR-11-2890

69. Liu X, Shi Y, Maag DX, et al. Iniparib nonselectively modifies cysteine-containing proteins in tumor cells and is not a bona fide PARP inhibitor. CLIN CANCER RES. 2012;18:510–523. doi:10.1158/1078-0432.CCR-11-1973

70. Mateo J, Ong M, Tan DS, Gonzalez MA, de Bono JS. Appraising iniparib, the PARP inhibitor that never was--what must we learn? NAT REV CLIN ONCOL. 2013;10:688–696. doi:10.1038/nrclinonc.2013.177

71. Venkitaraman AR. Cancer susceptibility and the functions of BRCA1 and BRCA2. CELL. 2002;108:171–182. doi:10.1016/s0092-8674(02)00615-3

72. Narod SA, Foulkes WD. BRCA1 and BRCA2: 1994 and beyond. NAT REV CANCER. 2004;4:665–676. doi:10.1038/nrc1431

73. Featherstone C, Jackson SP. DNA double-strand break repair. CURR BIOL. 1999;9:R759–R761. doi:10.1016/S0960-9822(00)80005-6

74. Wang JY. DNA damage and apoptosis. CELL DEATH DIFFER. 2001;8:1047–1048. doi:10.1038/sj.cdd.4400938

